# Cardiopulmonary resuscitation with Synchronized Ventilation: A new technique of Neonatal Resuscitation that reduces time to return of spontaneous circulation in a neonatal porcine model

**DOI:** 10.1101/2025.10.07.25337536

**Authors:** Shrieya Praveen, Megan O’Reilly, Raza Hyderi, Tze-Fun Lee, Marwa Ramsie, Georg M. Schmölzer

**Affiliations:** Centre for the Studies of Asphyxia and Resuscitation, Neonatal Research Unit, Royal Alexandra Hospital, Edmonton, Alberta, Canada; Department of Pediatrics, University of Alberta, Edmonton, Alberta, Canada

**Keywords:** NEWBORN, ASPHYXIA, CHEST COMPRESSION

## Abstract

**Introduction:** Current neonatal resuscitation guidelines recommend a 3:1 Compression:Ventilation ratio (3:1 C:V) during cardiopulmonary resuscitation. One of the concerns with 3:1 C:V is lung derecruitment, which contributes to a delay in achieving return to spontaneous circulation (ROSC). An alternative approach might be chest compression synchronized ventilation (CCSV), which delivers a ventilation with each chest compression thereby achieving lung recruitment. We hypothesized that in asphyxiated newborn piglets with cardiac arrest CCSV vs 3:1 C:V would decrease the time to ROSC.

**Methods:** Newborn piglets were anesthetized, intubated, instrumented, and exposed to 45-minute normocapnic hypoxia, followed by asphyxia. Piglets were randomized to either CCSV or 3:1 C:V. Piglets assigned to CCSV received 120 compressions/min with a ventilator-synchronized inflation delivered during every compression (CCSV-Mode, Weinmann Germany). In the 3:1 C:V group, piglets received 90 compressions/min and 30 ventilations/min. Compressions and ventilations were continued until ROSC. Continuous respiratory parameters, cardiac output, mean systemic artery pressures, and blood flows were measured.

**Results:** Sixteen neonatal mixed breed pigs (1-3 days of age, weighing 1.8-2.3kg) and were randomly assigned to CCSV or 3:1 C:V. The median (IQR) asphyxia time was not significantly different between CCSV (326 (275-405)sec) and 3:1 C:V (416 (266-475)sec) (p=0.442). Time to ROSC was significantly lower using CCSV with 68 (50-125)sec vs 170 (105-312)sec with 3:1 C:V (p=0.030). Rate of ROSC was 6/8 with CCSV and 5/8 with 3:1 C:V (p=1.000).

**Conclusions:** In a neonatal piglet model of asystolic cardiac arrest, CCSV resulted in a significantly faster time to ROSC compared to 3:1 C:V. Using CCSV might be an alternative to 3:1 C:V for neonatal resuscitation, but further studies are warranted.

## Introduction

Approximately 0.1%-1% of term infants require cardiopulmonary resuscitation (CPR) at birth primarily due to asphyxia, which necessitates a resuscitation approach that prioritizes ventilation. The current consensus on science and treatment recommendations recommend a 3:1 compression to ventilation ratio (3:1 C:V), which consists of 90 chest compressions (CC) and 30 ventilations per minute. However, the most effective CPR approach for newborn infants remains unknown.

Previous neonatal animal studies compared different C:V ratios including 2:1, 4:1, 9:3, and 15:2 and reported no difference in survival or time to return to spontaneous circulation times (ROSC)^1–3^. While all studies provided high quality CC, an overlooked effect is lung derecruitment, which occurs with every compression. Indeed, during the downward phase of compression, air is forced out of the lung, thereby resulting in lung derecruitment with reduced alveolar surface for gas exchange. This results in longer duration of CC, which is associated with increased risk of mortality and long-term neurological impairment.

Therefore, approaches that combine high quality CC with an improved ventilation strategy to prevent lung derecruitment might improve outcomes during neonatal CPR. Chest compression synchronized ventilation (CCSV), a novel resuscitation technique, may overcome these limitations of lung derecruitment. CCSV can be activated in the MEDUMAT Standard^2^ Ventilator (Weinmann Emergency Medical Technology, Hamburg, Germany) by pressing a button. In the CCSV mode, the flow sensor detects compression-induced expiratory flow (air forced out of the lungs) and the associated rise in airway pressure during the compression. Within ∼200–345 ms of the compression, the ventilator initiates an inflation during the down stroke of each CC with exhalation occurring during chest recoil.

This technique may maintain lung recruitment, improve gas exchange, and oxygenation. Previous studies using CCSV in adult piglets, reported conflicting results, with one study reporting improved partial pressure of arterial oxygen (PaO_2_) and lower partial pressure of arterial carbon dioxide (PaCO_2_) while another study reported no difference in either parameters^4–8^. As ventilation is the cornerstone of neonatal resuscitation, CCSV might improve outcomes compared to 3:1 C:V. We aimed to compared CCSV with 3:1 C:V in a piglet model of neonatal asphyxia. We hypothesized that in asphyxiated neonatal piglets CCSV compared to 3:1 C:V would reduce the time to ROSC.

## Methods

All experiments were conducted in accordance with the guidelines and approval of the Animal Care and Use Committee (Health Sciences), University of Alberta [AUP00002920], presented according to the ARRIVE 2.0 guidelines^9^, and registered at preclinicaltrials.eu (PCTE0000649). A graphical display of the study protocol is presented in Figure 1.

**Figure 1.**
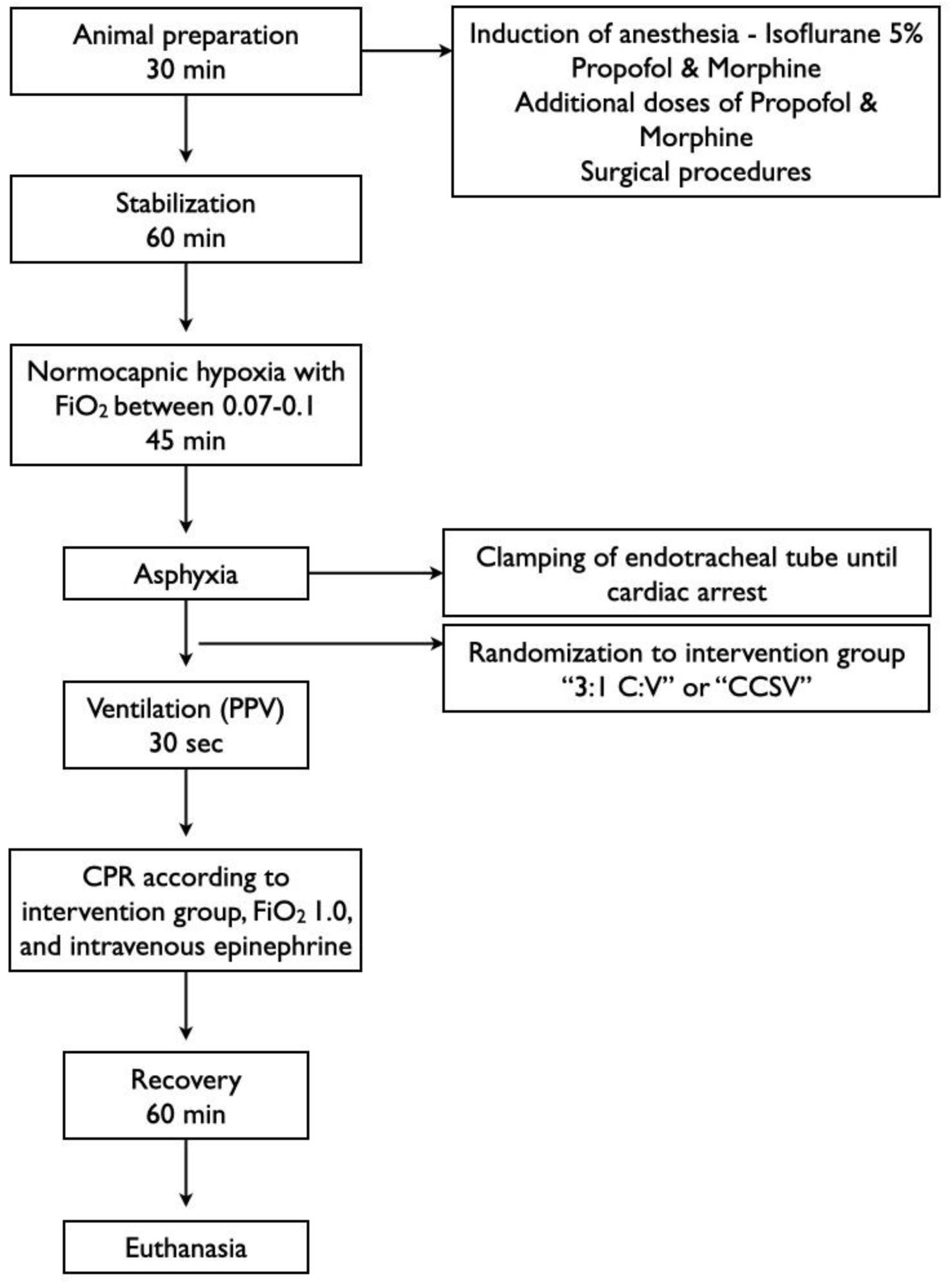
Study flow chart. PPV=positive pressure ventilation, CPR=cardiopulmonary resuscitation, CCSV=chest compression synchronized ventilation, 3:1 C:V= 3:1 compression to ventilation ratio, FiO_2_=fraction of inspired oxygen

### Inclusion and exclusion criteria

Neonatal mixed-breed piglets (1-3 days of age, weighing 1.7-2.8 kg) obtained on the day of experimentation from the University Swine Research Technology Centre were included. There were no exclusion criteria.

### Randomization

Piglets were randomly allocated to CCSV or 3:1 C:V. Randomization was block randomized with variable sized blocks using a 1:1 allocation using a computer-generated randomization program (https://www.randomizer.org). Sequentially numbered, sealed, brown envelopes containing the allocation were opened during the experiment.

### Blinding

Owing to the nature of the intervention, it was not feasible to blind the team to the allocated intervention. However, the randomization process enabled the concealment of the intervention until cardiac arrest was confirmed (by GMS). The statistical analysis was blinded to group allocation and only unblinded after statistical analysis was complete.

### Sample size and power estimates

Our primary outcome measure was time to ROSC. We previously reported that mean (SD) time to ROSC with 3:1 C:V was 388(83)sec^10^. We hypothesized that a sample size of 16 piglets (8 per group) would be sufficient to detect a 30% decrease in time to ROSC with 80% power and a 2-tailed alpha error of 0.05.

### Animal preparation

Piglets were instrumented as previously described with modifications^11,12^. Following induction of anesthesia using isoflurane, piglets underwent a tracheostomy for intubation. Pressure-controlled ventilation (Sechrist Infant Ventilator Model IV-100; Sechrist Industries, Anaheim, CA) was commenced at a respiratory rate of 16-20 breaths/min and pressure of 20/5cmH_2_O. Oxygen saturation was maintained within 90-100%, while glucose was provided via an intravenous infusion of 5% dextrose at 10mL/kg/hr. Intravenous propofol (5-10mg/kg/hr) and morphine (0.1mg/kg/hr) were provided to maintain anesthesia, and additional intravenous doses of propofol (1-2mg/kg) and morphine (0.05–0.1mg/kg) were administered as needed^11,12^. Piglet’s normothermic body temperature of 38.5–39.5°C was maintained with a heating pad and overhead warmer.

### Hemodynamic parameters

5-French Argyle® (Klein-Baker Medical Inc. San Antonio, TX) double-lumen and single-lumen catheters were inserted via the right femoral vein and artery, respectively. The femoral venous catheter was used for administration of fluids and medications and the arterial catheter for continuous arterial blood pressure monitoring in addition to arterial blood gas measurements. A real-time ultrasonic flow probe (2mm; Transonic Systems Inc., Ithica, NY) encircled the right common carotid artery to measure cerebral blood flow^11,12^.

Following surgical instrumentation, piglets were placed in the supine position and allowed to recover from surgical instrumentation until baseline hemodynamic measures were stable (minimum one hour). Ventilator rate was adjusted to maintain the partial arterial CO_2_ between 35-45mmHg, as determined by periodic arterial blood gas analysis. A Hewlett Packard 78833B monitor (Hewlett Packard Co., Palo Alto, CA) was used for the continuous measurement of heart rate, mean systemic arterial pressure, systemic systolic and diastolic arterial pressure, and percutaneous oxygen saturation throughout the experiment^11,12^.

### Respiratory Function Monitor

A respiratory function monitor (NM3, Respironics, Philips, Andover, MA) was used to continuously measure V_T_, airway pressures, and gas flow. The NM3 flow sensor has a fixed orifice pneumotach, which uses the pressure difference to calculate the gas flow passing through the sensor, which is then translated into the inspiratory and expiratory V_T_. The sensor was placed between the endotracheal tube and the ventilation device. V_T_ was calculated by integrating the flow signal^13^. The accuracy for gas flow is ±0.125 L/min^14^.

### Cerebral perfusion

Cerebral oxygenation (crSO_2_) was measured using the Invos^TM^ Cerebral/Somatic Oximeter Monitor (Invos 5100, Somanetics Corp., Troy, MI), which calculates crSO_2_ and expresses values as the percentage of oxygenated hemoglobin (oxygenated hemoglobin/total hemoglobin). The sensor was placed on the right forehead of the piglet and secured with wrap and tape. A slim cap provided light shielding. Regional oxygen saturation values were recorded every second at a sample rate of 0.13 Hz^15^.

### Experimental protocol

Following surgical instrumentation and stabilization, piglets were exposed to 45 min of normocapnic hypoxia, which was followed by asphyxia. Asphyxia was achieved by disconnecting the ventilator and clamping the endotracheal tube until cardiac arrest. Cardiac arrest was defined as zero carotid blood flow and no audible heartbeat during auscultation. A sequentially numbered, sealed brown envelope containing the allocation “CCSV” or “3:1 C:V” was opened.

Thirty seconds after confirmation of cardiac arrest, positive pressure ventilation (PPV) was provided for 30sec. If randomized to 3:1 C:V, PPV was provided with a Neopuff T-Piece (Fisher & Paykel, Auckland, New Zealand) with 100% oxygen, peak inspiratory pressure of 30cmH_2_O, positive end expiratory pressure of 5cmH_2_O, and gas flow of 10 L/min. If randomized to CCSV, PPV was provided with the MEDUMAT Standard^2^ ventilator (Weinmann Emergency Medical Technology, Hamburg, Germany) with 100% oxygen, peak inflation pressure of 30cmH_2_O, positive end expiratory pressure of 5cmH_2_O.

After 30sec of PPV, CC were started and were performed using the two-thumb CC technique at a rate of 120 compressions/min (guided by a metronome).

In randomized to 3:1 C:V, 90 compressions and 30 inflations per minute were given. Inflations were delivered using a T-Piece with a peak inflating pressure of 30cmH_2_O, a positive end-expiratory pressure of 5cmH_2_O, and 100% oxygen.

If randomized to CCSV, the CCSV mode on the MEDUMAT Standard^2^ ventilator was activated. In the CCSV mode, the ventilator synchronizes inflations with the down stroke of each chest compression. Synchronization is achieved when the ventilator’s sensors detect compression-induced expiratory flow (air forced out of the lungs) and the associated rise in airway pressure during the compression. The ventilator then delivers a positive-pressure inflation targeting a pre-set peak inflation pressure of 40 or 60 cmH_2_O, initiated within ∼200–345 ms of the compression. During chest recoil, exhalation occurs. Positive end-expiratory pressure is adjustable. A dedicated flow-trigger sensitivity control (1-5), used solely to detect compression-induced airflow, is adjustable to optimize synchronization and does not otherwise alter ventilator behaviour. The CCSV mode is designed for patients ≥10 kg. During the study, CCSV was delivered with a peak inflating pressure of 40 cmH_2_O, a positive end-expiratory pressure of 5 cmH_2_O, 100% oxygen. The trigger was adjusted between 1-5 to ensure optimized synchronization between ventilation and CC.

CCSV and 3:1 C:V were continued until ROSC or a maximum time resuscitation time of 10 min, whichever occurred first. Cardio-resuscitative drug, epinephrine, was administered (0.02mg/kg per dose) intravenously 2 min after the start of CC and thereafter every 3 min until ROSC, with a maximum of three doses. Each dose of epinephrine was followed by a 3 mL normal saline flush. ROSC was defined as an unassisted heart rate >100/min for at least 15 sec, detected by ECG. After ROSC, piglets recovered and were monitored for 60 min while continuously ventilated. Blood gases were collected at defined cardiac arrest, immediately after ROSC, and after 60 min of recovery. At the end of experimentation, piglets were euthanized with an intravenous overdose of sodium pentobarbital (120mg/kg).

### Tissue preparation and analysis

Tissue samples were only collected from piglets that survived one hour after ROSC. Following euthanasia, right lung tissue was collected. Lung tissue samples were snap frozen in liquid nitrogen and stored at −80 °C. Tissue samples were homogenized in lysis buffer (0.5 % Tween-20/PBS containing protease inhibitor cocktail), centrifuged (3,000xg for 10 min at 4 °C), the supernatants were collected, and protein concentration was quantified using the Bradford method. Evidence of lung injury was determined by quantification of the concentrations of the pro-inflammatory cytokines interleukin (IL)-6, and −8, and tumor necrosis factor −α in tissue homogenates by using commercially available ELISA kits as per manufacturer’s instructions (P6000B, P8000, PTA00; R&D Systems, Minneapolis, MN).

### Data collection and analysis

Demographics of study piglets were recorded. Transonic flow probes, heart rate, and pressure transducer outputs, and Millar catheter were digitized and recorded with LabChart^®^ programming software (ADInstruments, Houston, TX). Airway pressures, gas flow, tidal volume, and end-tidal CO_2_ were measured and analyzed using Flow Tool Physiologic Waveform Viewer (Philips Healthcare, Wallingford, CT). The data were tested for normality (Shapiro-Wilk and Kolmogorov-Smirnov test) and compared using Student’s t-test for parametric, Mann-Whitney U-test for nonparametric comparisons of continuous variables, and Fisher Exact-Test for categorical variables. The data are presented as mean (standard deviation-SD) for normally distributed continuous variables and median (interquartile range-IQR) when the distribution is skewed. *P*-values are 2-sided and *p*<0.05 was considered statistically significant. Statistical analyses were performed with SigmaPlot (Systat Software Inc, San Jose, CA).

## Results

Sixteen neonatal mixed breed pigs were obtained on the day of the experiment (1-3 days of age, weighing 1.8-2.3kg) and were randomly assigned to CCSV or 3:1 C:V. Characteristics at baseline are presented in Table 1. Blood gases at baseline, commencement of resuscitation, immediately after, and 60 min after ROSC are presented in Table 2.

**Table 1.** Baseline characteristics.

|  | **CCSV (n=8)** | **3:1 C:V (n=8)** | **p-value** |
| --- | --- | --- | --- |
| Age (days) | 2.5 (1.3-3.0) | 2.0 (2.0-3.0) | 0.798 |
| Weight (kg) | 1.9 (1.8-2.2) | 2.0 (1.8-2.3) | 0.652 |
| Gender (male/female) | 6/2 | 6/2 | 1.000 |
| Heart rate (bpm) | 160 (148-232) | 182 (175-207) | 0.693 |
| MAP (mmHg) | 72 (67-76) | 71 (64-76) | 0.900 |
| Carotid flow (mL/min) | 41 (33-55) | 54 (37-63) | 0.344 |
| Cerebral oxygenation (%) | 50 (43-52) | 50 (48-53) | 0.798 |
| pH | 7.49 (7.42-7.58) | 7.53 (7.40-7.61) | 0.959 |
| paCO<sub>2</sub> (torr) | 32 (29-35) | 31 (29-36) | 0.959 |
| paO<sub>2</sub> (torr) | 73 (63-85) | 68 (55-80) | 0.328 |
| Base excess (mmol/L) | 1.4 (-2.2~ 4.8) | 3.3 (-1.8~ 6.3) | 0.505 |
| Lactate (mmol/L) | 5.7 (4.6-6.0) | 4.9 (3.7-6.0) | 0.288 |

Data are presented as median (IQR); MAP=mean arterial blood pressure, PaCO<sub>2</sub>=partial pressure of arterial carbon dioxide, PaO<sub>2</sub>=partial pressure of arterial oxygen, CCSV=chest compression synchronized ventilation, 3:1 C:V= 3:1 compression to ventilation ratio

**Table 2.**
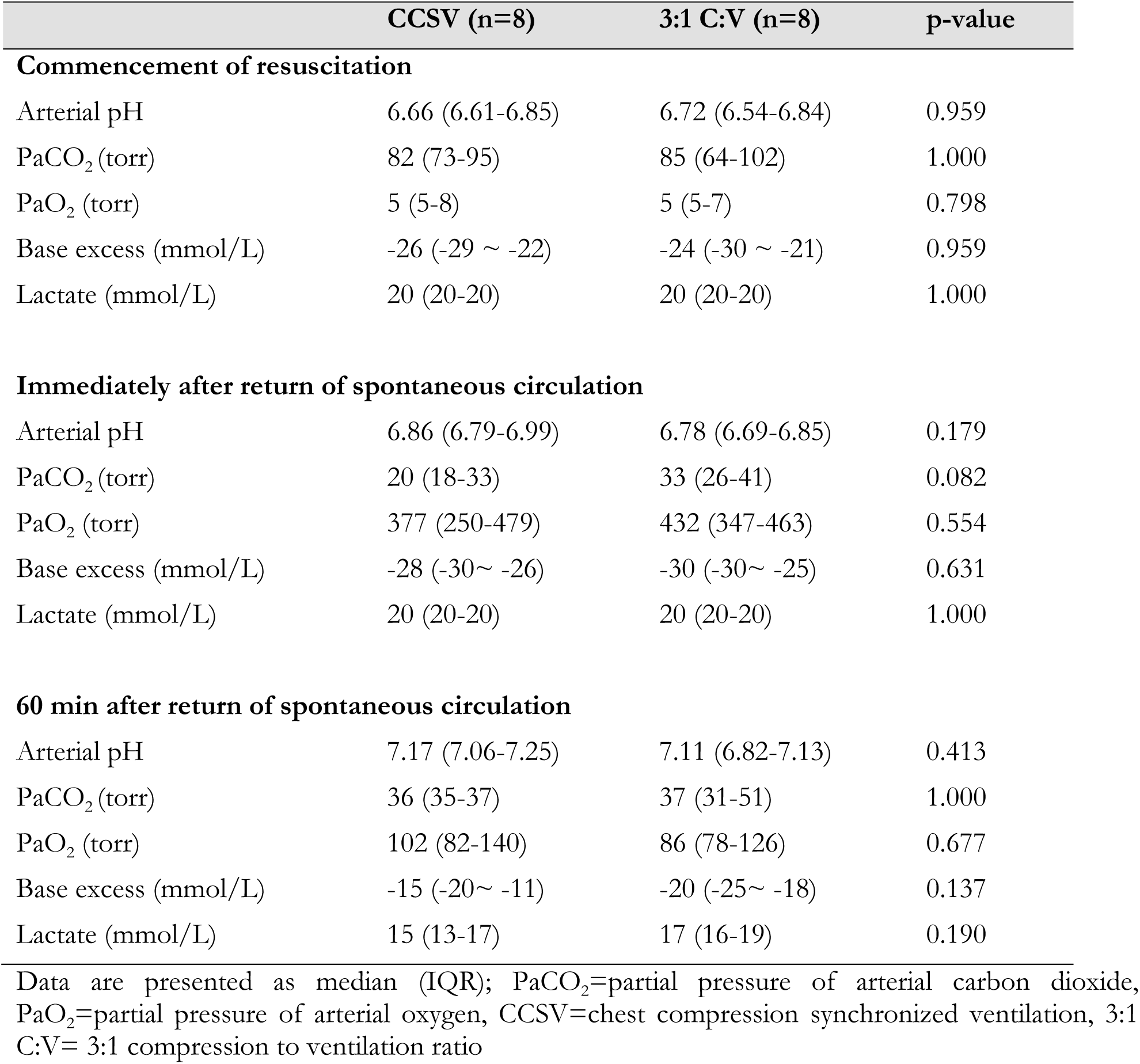
Blood gas changes throughout the experiment.

|  | **CCSV (n=8)** | **3:1 C:V (n=8)** | **p-value** |
| --- | --- | --- | --- |
| **Commencement of resuscitation** |  |  |  |
| Arterial pH | 6.66 (6.61-6.85) | 6.72 (6.54-6.84) | 0.959 |
| PaCO<sub>2</sub> (torr) | 82 (73-95) | 85 (64-102) | 1.000 |
| PaO<sub>2</sub> (torr) | 5 (5-8) | 5 (5-7) | 0.798 |
| Base excess (mmol/L) | -26 (-29 ~ -22) | -24 (-30 ~ -21) | 0.959 |
| Lactate (mmol/L) | 20 (20-20) | 20 (20-20) | 1.000 |
| **Immediately after return of spontaneous circulation** |  |  |  |
| Arterial pH | 6.86 (6.79-6.99) | 6.78 (6.69-6.85) | 0.179 |
| PaCO<sub>2</sub> (torr) | 20 (18-33) | 33 (26-41) | 0.082 |
| PaO<sub>2</sub> (torr) | 377 (250-479) | 432 (347-463) | 0.554 |
| Base excess (mmol/L) | -28 (-30~ -26) | -30 (-30~ -25) | 0.631 |
| Lactate (mmol/L) | 20 (20-20) | 20 (20-20) | 1.000 |
| **60 min after return of spontaneous circulation** |  |  |  |
| Arterial pH | 7.17 (7.06-7.25) | 7.11 (6.82-7.13) | 0.413 |
| PaCO<sub>2</sub> (torr) | 36 (35-37) | 37 (31-51) | 1.000 |
| PaO<sub>2</sub> (torr) | 102 (82-140) | 86 (78-126) | 0.677 |
| Base excess (mmol/L) | -15 (-20~ -11) | -20 (-25~ -18) | 0.137 |
| Lactate (mmol/L) | 15 (13-17) | 17 (16-19) | 0.190 |

Data are presented as median (IQR); PaCO<sub>2</sub>=partial pressure of arterial carbon dioxide, PaO<sub>2</sub>=partial pressure of arterial oxygen, CCSV=chest compression synchronized ventilation, 3:1 C:V= 3:1 compression to ventilation ratio

### Resuscitation and primary outcome

Median (IQR) time to asphyxia from endotracheal tube occlusion was 326 (275-405)sec and 416 (266-475)sec with CCSV and 3:1 C:V, respectively, p=0.442 (Table 3). Median time (IQR) to ROSC among survivors was significantly lower with CCSV with 68 (50-125)sec compared to 170 (105-312)sec with 3:1 C:V, p=0.030 (Table 3). The rate of ROSC with CCSV compared to 3:1 C:V was 6/8 (75%) vs. 5/8 (63%), respectively, p=1.000 (Table 3). The median (IQR) duration of resuscitation, which included non-survivors, with CCSV compared to 3:1 C:V was 97 (54-486)sec vs. 312 (133-600)sec, p=0.105, respectively (Table 3).

**Table 3.** Characteristics of asphyxia, resuscitation, and survival of asphyxiated piglets.

|  | **CCSV (n=8)** | **3:1 C:V (n=8)** | **p value** |
| --- | --- | --- | --- |
| Asphyxia time (sec) | 326 (275-405) | 416 (266-475) | 0.442 |
| ROSC time (sec) | 68 (50-125) | 170 (105-312) | 0.030 |
| Achieving ROSC <sup>†</sup> | 6 (75%) | 5 (63%) | 1.000 |
| CPR time (sec) | 97 (54-486) | 312 (133-600) | 0.105 |
| Epinephrine doses (n) | 0.5 (0-2.5) | 1 (1-3) | 0.328 |

Data are presented as median (IQR), unless indicated <sup>†</sup>n(%); CCSV=chest compression synchronized ventilation, 3:1 C:V= 3:1 compression to ventilation ratio, CPR=cardiopulmonary resuscitation, ROSC=return of spontaneous circulation.

### Changes in hemodynamic parameters

Hemodynamic parameters were similar at baseline, after asphyxiation, after ROSC, and throughout the 60 min post-ROSC observation period between groups (Figure 2).

**Figure 2:**
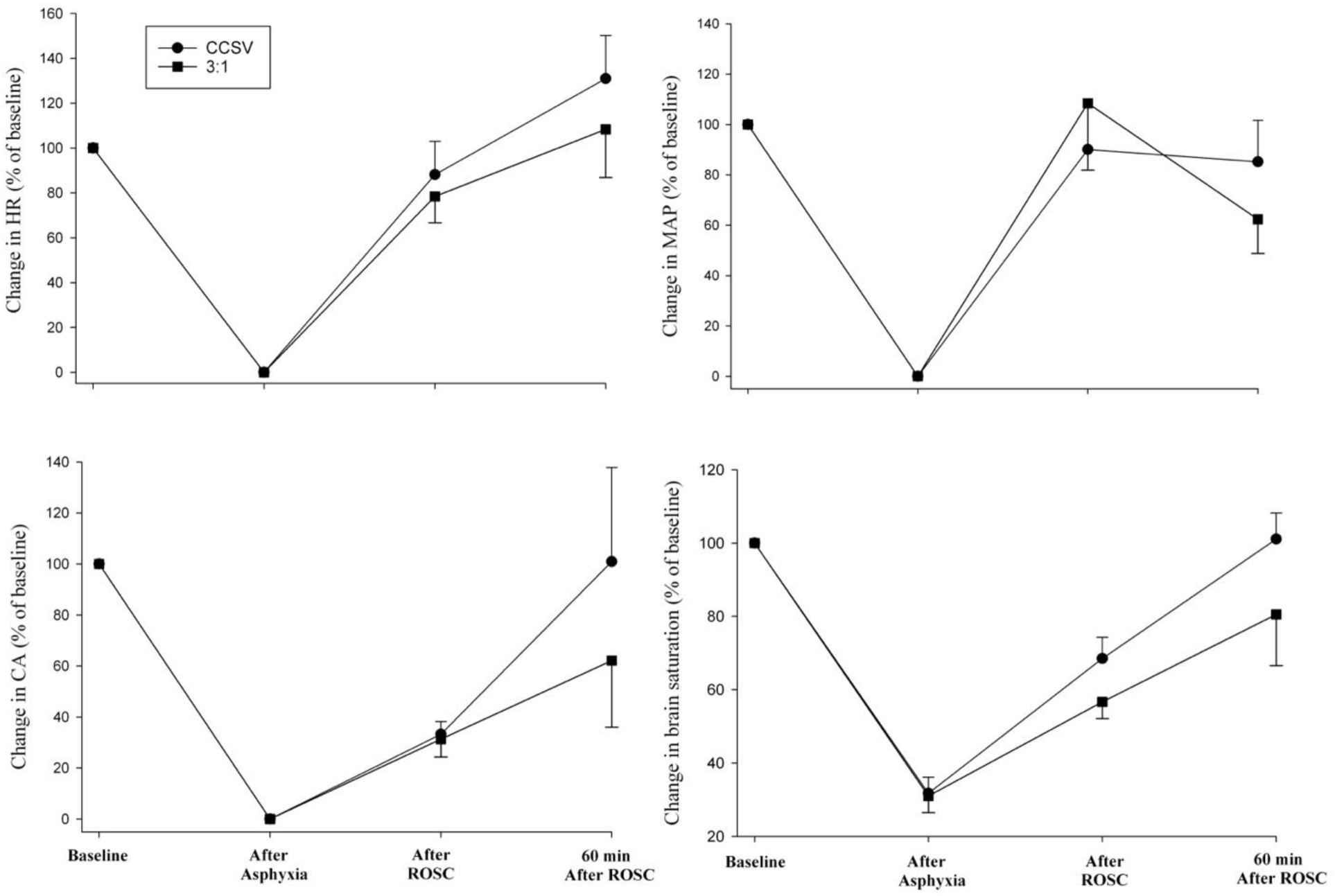
Hemodynamic Parameters. HR=heart rate, MAP=mean arterial pressure, CA=carotid artery flow

### Respiratory parameters

Respiratory parameters are presented in Table 4. Mean (SD) ventilation rate was 115(3.4) and 30(0) per minute with CCSV and 3:1 C:V (p=0.0002), respectively. Positive end expiratory pressure was 5(0.4) and 5 (0.6)cmH_2_O with CCSV and 3:1 C:V (p=0.5285), respectively. Mean (SD) minute ventilation was 555(164.2) mL/kg/min with CCSV and 188 (59.6)mL/kg/min with 3:1 C:V, (p=0.0002). Mean (SD) end-tidal CO_2_ was 14(6.8) vs. 5(5.3)mmHg (p=0.0166) with CCSV and 3:1 C:V, respectively.

**Table 4.**
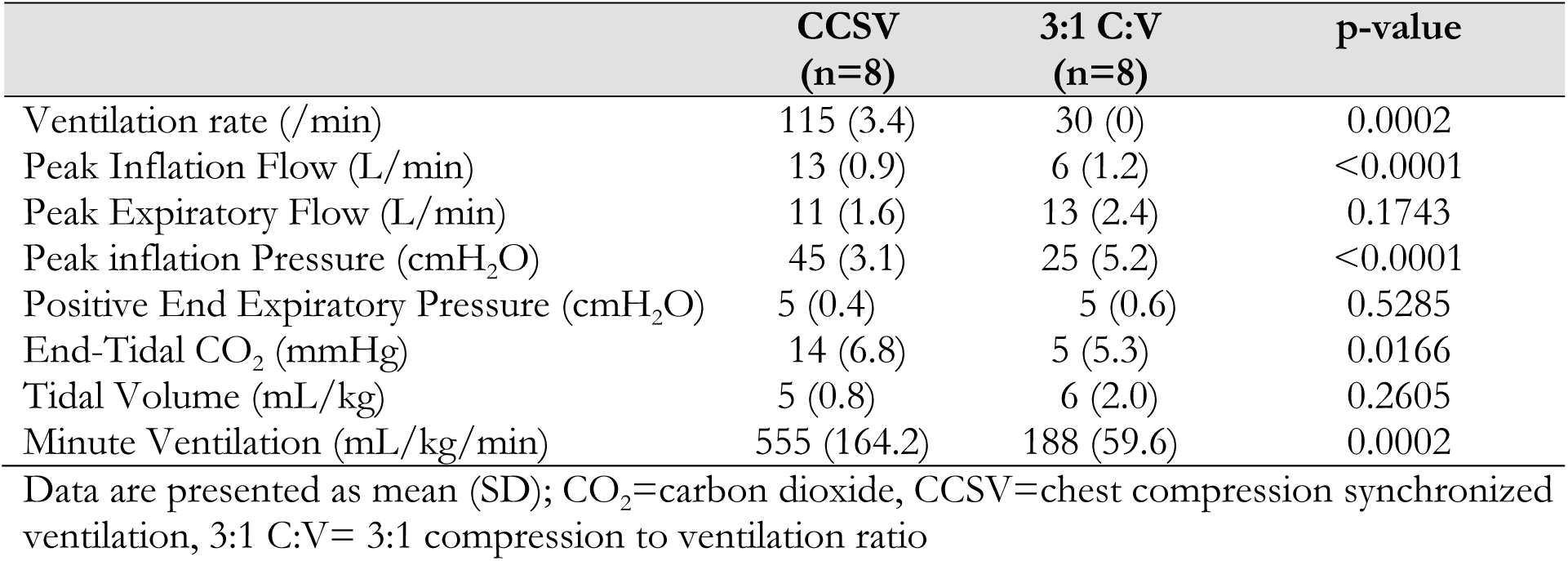
Respiratory parameters.

|  | **CCSV**<br>**(n=8)** | **3:1 C:V**<br>**(n=8)** | **p-value** |
| --- | --- | --- | --- |
| Ventilation rate (/min) | 115 (3.4) | 30 (0) | 0.0002 |
| Peak Inflation Flow (L/min) | 13 (0.9) | 6 (1.2) | <0.0001 |
| Peak Expiratory Flow (L/min) | 11 (1.6) | 13 (2.4) | 0.1743 |
| Peak inflation Pressure (cmH<sub>2</sub>O) | 45 (3.1) | 25 (5.2) | <0.0001 |
| Positive End Expiratory Pressure (cmH<sub>2</sub>O) | 5 (0.4) | 5 (0.6) | 0.5285 |
| End-Tidal CO<sub>2</sub> (mmHg) | 14 (6.8) | 5 (5.3) | 0.0166 |
| Tidal Volume (mL/kg) | 5 (0.8) | 6 (2.0) | 0.2605 |
| Minute Ventilation (mL/kg/min) | 555 (164.2) | 188 (59.6) | 0.0002 |

Data are presented as mean (SD); CO<sub>2</sub>=carbon dioxide, CCSV=chest compression synchronized ventilation, 3:1 C:V= 3:1 compression to ventilation ratio

### Lung inflammation

There was no difference in pro-inflammatory cytokines interleukin-6 and −8, and tumor necrosis factor-α (Figure 3).

**Figure 3:**
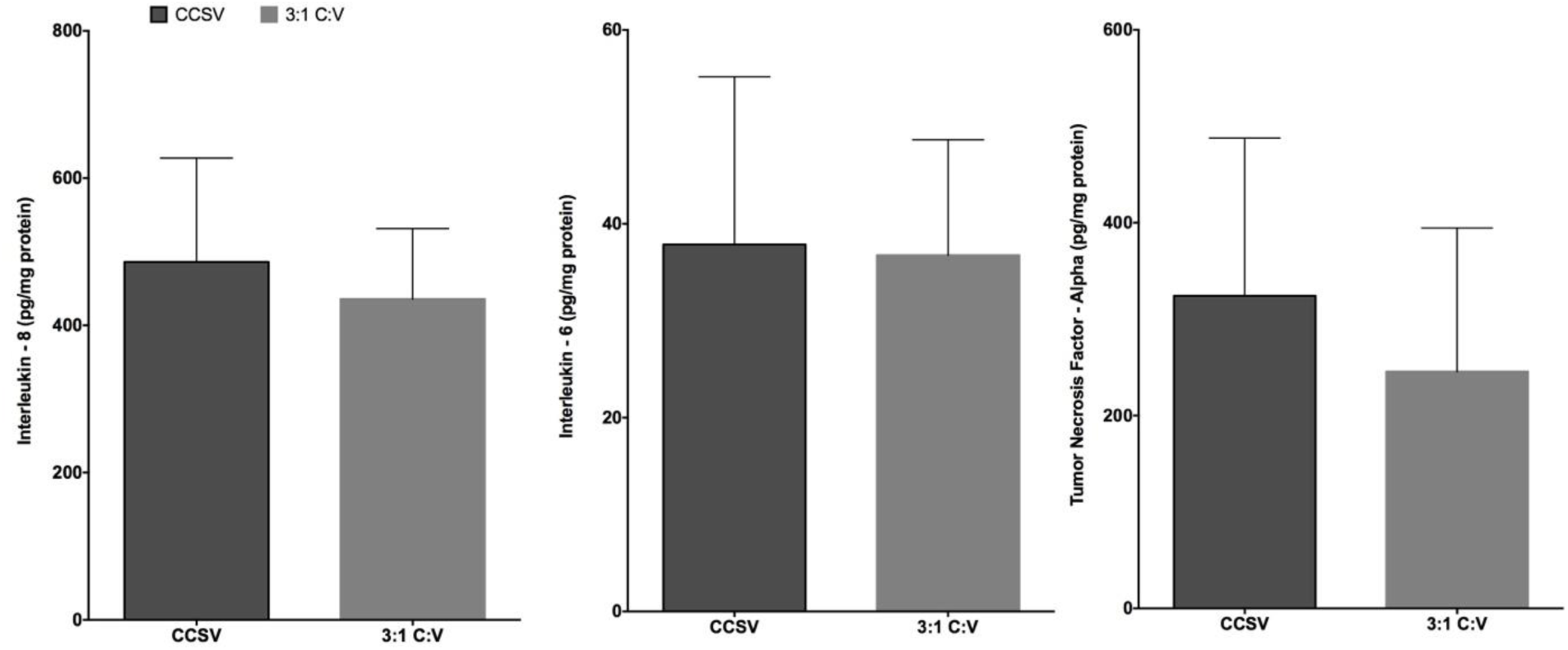
Interleukin (IL)-8, IL-6, and Tumor Necrosis Factor-alpha. CCSV=chest compression synchronized ventilation, 3:1 C:V= 3:1 compression to ventilation ratio

## Discussion

The primary cause for neonatal cardiac arrest is asphyxia, which occurs due to impaired gas exchange, leading to simultaneous hypoxia and hypercapnia^16–18^. To reverse asphyxia, the current neonatal consensus of science and treatment recommendations and resuscitation guidelines recommend a 3:1 C:V ratio. This approach combines CC, which generates blood flow to deliver oxygen to tissues^19–25^, and ventilations which aims to reverse hypoxia and hypercapnia^26,27^. However, animal studies have highlighted that CC causes lung derecruitment. With every compression, air is forced out of the lung. We have shown that with every 3:1 C:V cycle of 3 compressions and 1 inflation, there is a net lung volume loss^28^. Lung derecruitment during CPR is associated with longer time to ROSC due to the poorer gas exchange. A longer time to ROSC is further associated with significant increase in mortality long-term neurological and health impairment^29–32^. Therefore, it is prudent to improve ventilation strategies during CPR to allow for quicker reversal of hypoxia and hypercapnia^16–18^.

CCSV is a novel approach in which a ventilator flow sensor detects the compression-induced expiratory air flow and triggers an inflation during the downward movement of the chest with exhalation during chest recoil. To our knowledge, this is the first study examining CCSV in a survival piglet model in newborns, children, or adults. The results of this study can be summarized as follows: i) CCSV resulted in a significantly faster time to ROSC compared to 3:1 C:V, ii) end-tidal CO_2_ and minute ventilation was significantly improved, while iii) hemodynamic parameters and venous return was not different between groups.

During 3:1 C:V ratio, compressions are paused after every third compression to deliver one inflation, a pattern expected to transiently lower intrathoracic pressure. The purpose of inflations during CPR is to deliver adequate tidal volume to support gas exchange; each inflation also raises intrathoracic pressure and augments antegrade blood flow. Animal studies have described lung derecruitment during CC. In an adult pig, atelectasis increased during chest compression only CPR from 25% pre-CPR to 75% post-CPR. We also described lung derecruitment in neonatal CPR with up to 4.5 mL of volume lost during the 3 compressions between inflations. In our study, CCSV delivered a similar tidal volume to 3:1 C:V, however due to the higher ventilation rate, the minute ventilation was significantly improved, which contributed to the faster time to ROSC. Whereas current guidelines recommend 30 inflations per minute, CCSV used 120 inflations per minute, a fourfold increase that substantially raised minute ventilation. Consistent with this, exhaled CO_2_ was significantly higher with CCSV, indicating increased alveolar ventilation, pulmonary perfusion (right cardiac output), and metabolic CO_2_.

Across five studies spanning adult swine models and a small clinical trial, CCSV shows mixed results^4–8^. In a 24-pig trial, CCSV produced higher PaO_2_, lower PaCO_2_, higher mean arterial pressure, and near-normal mixed-venous pH versus intermittent positive-pressure ventilation (IPPV) or bilevel ventilation, while ROSC rates were similar (IPPV 5/8, bilevel 6/8, CCSV 4/7)^5^. A prolonged arrest swine model comparing 30:2 with continuous compressions plus asynchronous ventilation found no significant differences in gas exchange or hemodynamics^6^. A crossover porcine study confirmed that multiple CCSV presets raised PaO_2_ and prevented arterial-pressure decline relative to IPPV^4^. Importantly, in swine, combining CCSV with aortic balloon occlusion increased perfusion metrics, yielded ROSC in 7/7 animals, and improved post-resuscitation organ outcomes versus IPPV^8^. In contrast, a randomized pilot in out-of-hospital cardiac arrest patients showed within-group improvements with CCSV but no superiority over IPPV between groups^7^. We found similar results in our study, with PaCO_2_ values as low as 20mmHg with CCSV. Low PaCO_2_ values have been correlated with periventricular leukomalacia and worse neurological outcomes in preterm infants at 18 months^33^.

## Limitations

Our use of a piglet asphyxia model is a considerable strength of this translational study, because this model closely mimics delivery room events with a gradual onset of severe asphyxia leading to bradycardia. However, several limitations should be considered before general application of CCSV in future clinical neonatal resuscitation trials. Our model uses piglets that have undergone the fetus to neonate transition and do not possess fetal features (i.e., no lung fluid present or a patent ductus arteriosus). Additionally, our model uses piglets that were sedated/anesthetized, and intubated with a tightly sealed endotracheal tube to prevent leak, which may not occur in the delivery room. Nevertheless, our findings remain relevant despite these limitations, because several previous studies with our post-transitional model have been successfully translated into neonatal clinical randomized trials.

## Conclusions

CCSV during CPR in newborn piglets significantly improved ROSC in a porcine model of neonatal resuscitation. This is of considerable clinical relevance, because improved respiratory and hemodynamic parameters potentially minimize morbidity and mortality in newborn infants.

## Acknowledgment

We would like to thank Weinmann Emergency Medical Technology for providing the MEDUMAT Standard2 ventilator for this study.

## Disclosures

### Data availability

The datasets generated and analyzed for this study are available from the corresponding author (GMS), upon reasonable request.

### Author’s contribution

Conception and design: GMS, MOR, TFL

Collection and assembly of data: GMS, MOR, MR, TFL, RH, SP

Analysis and interpretation of the data: GMS, MOR, MR, TFL, RH, SP

Drafting of the 1^st^ draft: SP

Critical revision of the article for important intellectual content: GMS, MOR, MR, TFL, RH, SP

Final approval of the article: GMS, MOR, MR, TFL, RH, SP

